# Clinical symptoms among ambulatory patients tested for SARS-CoV-2

**DOI:** 10.1101/2020.10.20.20213272

**Authors:** Jessie R. Chung, Sara S. Kim, Michael L. Jackson, Lisa A. Jackson, Edward A. Belongia, Jennifer P. King, Richard K. Zimmerman, Mary Patricia Nowalk, Emily T. Martin, Arnold S. Monto, Manjusha Gaglani, Michael E. Smith, Manish Patel, Brendan Flannery

**Author notes:** Corresponding author: Jessie R. Chung, Influenza Division, Centers for Disease Control and Prevention, 1600 Clifton Rd NE, Mailstop 24/7, Atlanta, Georgia, 30329. Conflict of Interest Disclosures: The authors have no financial relationships relevant to this article to disclose.

## Abstract

We compared symptoms and characteristics of 4961 ambulatory patients with and without laboratory-confirmed SARS-CoV-2 infection. Findings indicate that clinical symptoms alone would be insufficient to distinguish between COVID-19 and other respiratory infections (e.g., influenza) and/or to evaluate the effects of preventive interventions (e.g., vaccinations).

## Introduction

Since early 2020, pre-established influenza surveillance systems have contributed substantially to monitoring the coronavirus disease 2019 (COVID-19) pandemic in the United States. For syndromic surveillance, COVID-like illness is defined as acute illness with fever and either cough or shortness of breath, or a COVID-19 diagnostic code. To evaluate effectiveness of interventions to prevent mild illness associated with SARS-CoV-2 infection, standardized clinical criteria in addition to laboratory confirmation of infection will be needed. In March 2020, participating sites in the US Influenza Vaccine Effectiveness (Flu VE) research network revised study inclusion criteria to prospectively enroll persons with acute febrile or respiratory illness, characterized by reported fever or cough or shortness of breath, who had been tested for SARS-CoV-2 infection. Here we compare symptoms and characteristics of persons with and without laboratory-confirmed SARS-CoV-2 infection.

## Methods

At the study sites of the US Flu VE Network in Michigan, Pennsylvania, Texas, Washington, and Wisconsin, research staff screened for study eligibility among persons of all ages who had sought medical care (e.g., telehealth, primary care, urgent care, and emergency departments) and/or COVID-19 testing for an acute respiratory illness. Eligible individuals reported acute illness with fever/feverishness, cough, or shortness of breath/difficulty breathing and had a respiratory specimen collected for SARS-CoV-2 testing within 10 days of illness onset. At four sites, research staff contacted and screened patients for eligibility by telephone; at one site (Pennsylvania), a sample of potentially eligible patients was screened for eligibility by phone or online survey [4]. Standardized questionnaires collected demographic information, general health status, health-related behaviors such as cigarette smoking or vaping, healthcare-related employment, and contact with a person with confirmed COVID-19. Questionnaires also solicited information about twelve specific symptoms experienced since illness onset (Figure).

Respiratory specimens were tested for SARS-CoV-2 RNA using reverse-transcription polymerase chain reaction (RT-PCR) assays at study sites. Days from illness onset to testing were calculated using clinical specimen collection date. We classified persons with and without COVID-19 based on SARS-CoV-2 RT-PCR test results; persons tested by antigen detection assays alone were excluded. Underlying medical conditions, associated with increased risk of severe illness from COVID-19 (e.g., cancer, diabetes, chronic obstructive pulmonary disease, or hypertension) [5], were obtained for participants at four study sites from electronic medical records based on International Classification of Diseases (ICD)-10 diagnostic codes from medical encounters during the past year, as previously described [6]. Comparisons between persons with and without COVID-19 were made using chi-square tests (categorical variables) or Wilcoxon rank-sum tests (continuous variables). The p-value for significance was set at 0.05.

This study was approved by institutional review boards at the Centers for Disease Control and Prevention and all participating sites.

## Results

Between March 26 and August 15, 2020, 5760 patients were interviewed and 4961 were included in analyses. Of 799 patients excluded, 402 (50%) were swabbed, tested or interviewed more than 10 days after symptom onset, 31 (4%) had inconclusive RT-PCR results or had a SARS-CoV-2 antigen test (n=11), 82 (10%) were asymptomatic prior to enrollment, and 284 (36%) did not meet clinical criteria during the enrollment interview (i.e., no reported cough, fever/feverishness or shortness of breath/difficulty breathing). Of 284 not meeting clinical criteria, 50 (18%) reported decreased taste or smell.

Of 4961 included, 916 persons with laboratory-confirmed COVID-19 and 4045 with symptoms but without COVID-19 were included. Patient age and race/ethnicity differed by case status (p<0.01 for both, Supplemental Table). Compared with persons without COVID-19, persons with COVID-19 were more likely to be adults aged 18–49 years (65% vs 56%) and less likely to be younger than 18 years (6% vs 14%). Persons with COVID-19 more frequently self-identified as Hispanic/Latino (16% vs 7%) or non-Hispanic/Latino Black or African American (8% vs 6%) and less frequently as non-Hispanic/Latino White or Caucasian (70% vs 81%). Among patients with information on underlying chronic medical conditions, persons with COVID-19 less frequently had documented underlying medical conditions than persons without COVID-19 (42% vs 47%; p=0.02). Among adults aged ≥18 years, similar percentages of persons with and without COVID-19 identified as healthcare workers (25% vs 26%; p=0.62); persons with COVID-19 less frequently reported cigarette smoking or vaping (13% vs. 21%, p<0.01). During the 14 days before symptom onset, contact with a person with confirmed COVID-19 was reported by 59% of persons with COVID-19 versus 18% of persons without COVID-19 (p<0.01).

**Table.** Characteristics of 4961 ambulatory patients with acute respiratory illness clinically tested for SARS-CoV-2 by RT-PCR; US Influenza Vaccine Effectiveness Study, March 23 – August 15, 2020.

| Characteristic | Persons with COVID-19 |  | Persons without COVID-19 |  | P-value |
| --- | --- | --- | --- | --- | --- |
|  | N | Column % | N | Column % |  |
| **Total** | 916 | 100 | 4045 | 100 |  |
| **Study site** |  |  |  |  | <0.01 |
| Michigan | 32 | 3 | 269 | 7 |  |
| Pennsylvania | 330 | 36 | 1056 | 26 |  |
| Texas | 272 | 30 | 755 | 19 |  |
| Washington | 22 | 2 | 586 | 14 |  |
| Wisconsin | 260 | 28 | 1379 | 34 |  |
| **Age group (years)** |  |  |  |  | <0.01 |
| ≤4 | 12 | 1 | 227 | 6 |  |
| 5-17 | 40 | 4 | 332 | 8 |  |
| 18-49 | 591 | 65 | 2250 | 56 |  |
| 50-64 | 192 | 21 | 891 | 22 |  |
| ≥65 | 81 | 9 | 345 | 9 |  |
| **Female** | 355 | 39 | 1338 | 33 | <0.01 |
| **Race<sup>1</sup>** |  |  |  |  |  |
| White or Caucasian | 712 | 78 | 3514 | 87 | <0.01 |
| Black or African American | 95 | 10 | 286 | 7 | <0.01 |
| Asian | 25 | 3 | 119 | 3 | 0.73 |
| Native Hawaiian or other Pacific Islander | 2 | 0.2 | 18 | 0.5 | 0.33 |
| American Indian or Alaska Native | 9 | 1 | 76 | 2 | 0.06 |
| Other race | 98 | 11 | 179 | 4 | <0.01 |
| **Ethnicity<sup>2</sup>** |  |  |  |  |  |
| Hispanic or Latino | 144 | 16 | 285 | 7 | <0.01 |
| **Race and Ethnicity<sup>3</sup>** |  |  |  |  | <0.01 |
| Non-Hispanic or Latino, White or Caucasian | 642 | 70 | 3252 | 81 |  |
| Non-Hispanic or Latino, Black or African American | 76 | 8 | 231 | 6 |  |
| Non-Hispanic or Latino, other race | 51 | 6 | 261 | 6 |  |
| Hispanic or Latino, any race | 144 | 16 | 285 | 7 |  |
| **Healthcare worker<sup>4</sup>** | 218 | 25 | 907 | 26 | 0.62 |
| **Contact with confirmed person with COVID-19<sup>5</sup>** | 451 | 59 | 601 | 18 | <0.01 |

<sup>1</sup> Participants could select more than one category. Race was missing for 6 (<1%) participants.
<sup>2</sup> Missing for 19 (<1%) participants.
<sup>3</sup> Missing for 19 (<1%) participants.
<sup>4</sup> Only asked of participants aged ≥18 years. Denominators were 861 (3 adults did not answer) for persons with COVID-19 and 3470 (16 adults did not answer) for persons without COVID-19.
<sup>5</sup> Missing (i.e., responded “don’t know” or refused to answer) for 808 participants including 153 persons with and 655 persons without COVID-19.

Overall, patients reported a median of 3 days (interquartile range [IQR], 2–5) of symptoms before specimen collection for diagnostic SARS-CoV-2 testing, and a median of 6 days (IQR, 4– 8) of symptoms at the time of interview. Time between symptom onset and specimen collection did not differ between persons with and without COVID-19 (p=0.17). Among qualifying symptoms (fever/feverishness, cough or shortness of breath/difficulty breathing), cough was most commonly reported (86% of persons with and 83% of persons without COVID-19; p<0.01) (Figure). Shortness of breath/difficulty breathing was reported less frequently by persons with COVID-19 than without (40% versus 47%; p<0.01). Overall, 4898 (99%) patients meeting clinical screening criteria for inclusion in this research study reported fever/feverishness and/or cough. Persons with COVID-19 reported a median of 7 (IQR, 5–8) of the assessed symptoms versus 6 (IQR, 4–7) for persons without COVID-19 (p<0.01). Among 4102 participants asked, 59% of persons with COVID-19 versus 19% of persons without COVID-19 reported diminished taste or smell (p<0.01). Generalized symptoms (muscle aches or headache) and gastrointestinal symptoms (vomiting, diarrhea, or abdominal pain) were more common among persons with COVID-19 (91% and 57%, respectively) than among those without COVID-19 (83% and 50%, respectively) (p<0.01 for both).

**Figure.**
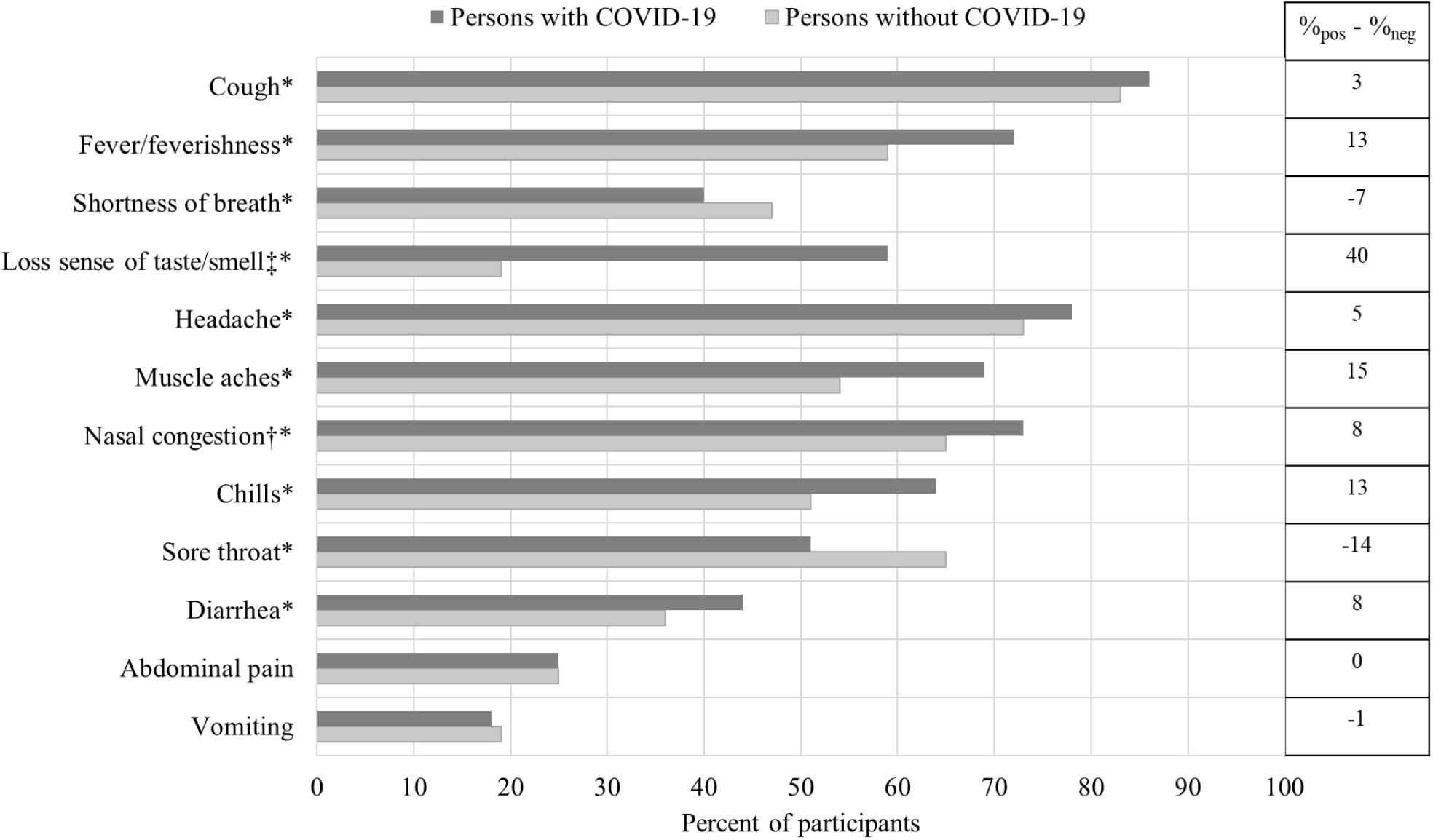
Self-reported presence of clinical symptoms among 4961 ambulatory persons with acute respiratory illness, by clinical SARS-CoV-2 RT-PCR test result, US Influenza Vaccine Effectiveness Network, March 23–August 15, 2020. * Indicates statistically significant difference between persons with and without COVID-19 (i.e., p<0.05). ‡ A subset of participants were asked about loss or decreased sense of taste/smell including 850 persons with COVID-19 and 3252 persons without COVID-19. † A subset of participants were asked about nasal congestion including 613 persons with COVID-19 and 2687 persons without COVID-19.

## Discussion

For control of the COVID-19 pandemic, the Centers for Disease Control and Prevention (CDC) recommends testing symptomatic persons when there is a concern of potential COVID-19 [2]. Clinical signs and symptoms consistent with COVID-19 are common among patients seeking healthcare for mild respiratory illness and may not inform decisions on who should be tested for SARS-CoV-2 infection. However, future evaluation of effectiveness of interventions, including treatment and vaccination, for prevention of illness or progression to severe disease will require systematic testing of medically attended illnesses meeting standard clinical criteria [3]. In this study conducted in five U.S. locations, persons with mild illness seeking clinical care or COVID-19 testing who met simple, standardized screening criteria (reported fever, cough, or shortness of breath of *≤*10 day duration) commonly reported other respiratory, gastrointestinal, or systemic symptoms. Although persons with COVID-19 were more likely than those without COVID-19 to report gastrointestinal symptoms (vomiting, diarrhea, or abdominal pain) or other symptoms such as muscle aches or headache, the largest difference was observed in diminished or complete loss of taste or smell, reported by more than half of persons with laboratory-confirmed COVID-19 versus one in five persons without COVID-19. Because of the wide overlap in COVID-19 symptoms with those of other respiratory illnesses, laboratory confirmation of SARS-CoV-2 infection will be critical, not only for limiting disease spread, contact tracing, and monitoring clinical course, but also for assessing the effectiveness of interventions during periods of co-circulation of SARS-CoV-2 and other respiratory viruses, including influenza.

Rapid identification of SARS-CoV-2 infection among symptomatic patients seeking healthcare is critical to limit disease spread. Participants in this study, including symptomatic healthcare workers and patients reporting contact with a person with confirmed COVID-19, reported a median of 3 days of illness before collection of respiratory specimens for viral testing; 25% reported ≥5 days. Delays in seeking healthcare may result in disease transmission without strict adherence to recommended preventive measures, especially among persons with a low index of suspicion for COVID-19. While contact with a known COVID-19 case was strongly associated with SARS-CoV-2 infection, almost half of people with COVID-19 did not report exposure from another person with known COVID-19.

One limitation of this analysis is that only patients meeting screening criteria were included, thus sensitivity and specificity of other research case definitions were not assessed. In addition, symptom lists at some study sites did not assess symptoms commonly reported by outpatients with confirmed COVID-19 (such as fatigue). Because enrollment in this research study occurred after a clinical swab was obtained for testing, some patients might have been aware of their test results at the time of enrollment and symptom assessment, which could have influenced responses to some questions. Finally, persons without COVID-19 (i.e., negative for SARS CoV-2 by RT-PCR) may have had false negative test results; however, to reduce misclassification, cases were classified based on results of molecular assays, and participants were tested within 10 days of illness onset.

In addition to establishing clinical criteria for mild illness to evaluate interventions to prevent COVID-19, systematic testing for SARS-CoV-2 and influenza will be needed when both viruses circulate for assessing effectiveness of 2020–2021 seasonal influenza vaccines [4, 9]. Symptom criteria that can be rapidly assessed before enrollment in research studies of influenza and COVID-19 would help ensure systematic testing for both viruses. Sensitivity and specificity of screening criteria including subjective fever, cough, and loss of taste or smell should be assessed among ambulatory persons with COVID-19 for use in epidemiologic studies [9].

## Supporting information

STROBE checklist for case-control studies

## Data Availability

Data may be made available in accordance with CDC data availability policy.

## Disclaimer

The findings and conclusions are those of the authors and do not necessarily represent the official position of the Centers for Disease Control and Prevention.

## Acknowledgments

Elizabeth Armagost, Deanna Cole, Terry Foss, Hannah Gourdoux, Erica Graves, Kayla Hanson, Linda Heeren, Lynn Ivacic, Jacob Johnston, Julie Karl, Diane Kohnhorst, Erik Kronholm, Karen McGreevey, Huong McLean, Jennifer Meece, Vicki Moon, Rebecca Pilsner, DeeAnn Polacek, Martha Presson, Carla Rottscheit, Jackie Salzwedel, Julian Savu, Sandy Strey, Melissa Wendt, and Gail Weinand from Marshfield Clinic Research Institute; Theresa M. Sax, Klancie Dauer, GK Balasubramani, Leah McKown, Emily Welsh, Michael Susick, Andrew Ho, and Meredith Axe from University of Pittsburgh; Joshua G. Petrie, Lois E. Lamerato, Ryan E. Malosh, E.J. McSpadden, Hannah Segaloff, Caroline K.Cheng, Rachel Truscon, Emileigh Johnson, Armanda Kimberly, Anne Kaniclides, Amy Getz, Kim Beney, Sarah Bauer, Michelle Groesbeck, Joelle Baxter, Rebecca Fong, Sarah Davenport, Miranda Viars, Micah Wildes, Regina Lehmann-Wandell, Kayla Morse, Rachel Phillips, Nicole Hermes from University of Michigan, Ann Arbor, and Henry Ford Health System, Detroit, Michigan; C. Hallie Phillips, Erika Kiniry, Stacie Wellwood, Brianna Wickersham, Suzie Park, Matt Nguyen, Rachael Burganowski from Kaiser Permanente Washington Health Research Institute; Kayan Dunnigan, Kempapura Murthy, Chandni Raiyani, Marcus Volz, Kimberley Walker, Martha Zayed, Mary Kylberg, Jeremy Ray, Deborah Price, Natalie Settele, Jennifer Thomas, Jaime Walkowiak, Michael Reis, Madhava Beeram, Arundhati Rao and Alejandro Arroliga from Baylor Scott & White Health

